# A Second Wave? What Do People Mean By COVID Waves? – A Working Definition of Epidemic Waves

**DOI:** 10.1101/2021.02.21.21252147

**Authors:** Stephen X. Zhang, Francisco Arroyo Marioli, Renfei Gao

## Abstract

Policymakers and researchers describe the COVID-19 epidemics by waves without a common vocabulary on what constitutes an epidemic wave, either in terms of a working definition or operationalization, causing inconsistencies and confusions. A working definition and operationalization can be helpful to characterize and communicate about epidemics. We propose a working definition of epidemic waves in the ongoing COVID-19 pandemic and an operationalization based on the public data of the effective reproduction number *R*. Our operationalization characterizes the numbers and durations of waves (upward and downward) in 179 countries and reveals patterns that can enable healthcare organizations and policymakers to make better description and assessment of the COVID crisis to make more informed resource planning, mobilization, and allocation temporally in the continued COVID-19 pandemic.

**ONE SENTENCE SUMMARY:** A working definition and operationalization of waves to enable common ground to understand and communicate COVID-19 crisis.

---

On June 16, 2020, Mike Pence, US vice president then, penned an op-ed in The Wall Street Journal headlined: “There isn’t a coronavirus ‘second wave’” (1). Yet on the same day, the American top federal infectious disease expert, Dr. Anthony Fauci, countered via The Wall Street Journal that: “People keep talking about a second wave,” he said. “We’re still in a first wave” (2). A Google web search revealed over 30 million mentions of COVID “second waves” by early Nov 2020. What do they mean by “first wave” and “second wave”? Unfortunately, “a wave is just a metaphor … -a term without a precise definition in epidemiology” (3).

Despite its lack of a definition, the concept of second wave has gained huge popularity, with broad implications. For instance, the UK Prime Minister warned all UK citizens that they must “prepare for the second wave of COVID” (4). Israeli authorities released officially that “the country was going through a second wave” (3). And the Chief Medical Officer of Australia, Brendan Murphy, said a second wave would be “the most worrying thing of all”. Similarly, scientists have used the term repeatedly. For example, A Lancet article has warned about the possibility of a second wave of COVID-19 pandemic as early as April 2020 (5). All of us want to know whether their places are experiencing the first wave, have gone through the peak of it, or are battling the second wave, third round, etc, because such assessment of the COVID epidemics carries profound implications on people’s lives, works, and physical and mental health (6). After all, the concept of waves in the past pandemics, such as the 1918 influenza epidemic (7), greatly helped people to understand those pandemics in greater detail to enable better communication, policies and actions

The article aims to bring a working definition of what constitutes a wave in the ongoing COVID pandemic so that people can have a common language to use, interpret and discuss epidemic waves, which are increasingly a concern worldwide. The working definition does not aim to offer a strict definition of waves in epidemiology but instead uses the prevailing characterizations of epidemic waves to reduce the arbitrariness that muddles the tumultuous concept at present. A thorough search of the academic literature and the popular media reveals M.D., epidemiologists, and policymakers do share the same underlying characterization of the term wave to give it some conceptual clarity. Popular media state “the word ‘wave’ implies a natural pattern of peaks and valleys” (8). Similarly, WHO, clarified “in order to say one wave is ended, the virus has to be brought under control and cases have to fall substantially,… Then for a second wave to start, you need a sustained rise in infections” (9). “A wave is also different to a “spike” in cases — another frequently used term” (3). “A spike [or upsurge] is a momentary phenomenon” (3). Moreover, in the case of epidemic waves of COVID, “it is probably not realistic for the number of new cases to drop to zero, but ideally one would like to see sustained decreases in the number of new cases over time or stability in the number of new cases over time” (10).

Such convergence in conceptual clarity of the term “wave” allows us to offer a working definition of the concept based on two key defining characteristics: 1) an epidemic wave constitutes some upward and/or downward periods; 2) the increase in an upward period or the decrease in a downward period have to be substantial by sustaining over a period of time to distinguish them from an uptick, a downtick, reporting errors, or volatility in new cases.

We propose to operationalize the epidemic wave in the ongoing COVID-19 pandemic by a publicly available popular real-time index, the effective reproduction number, *R*, which refers to the average number of people infected by a single infectious individual in real-time as the epidemic happens (11). When R>1, the number of cases increases, and when R<1 the number of cases decreases. *R* plays a central role in the epidemiology of infectious diseases and the *R* values are, because *R* estimates are not sensitive to potential model misspecification and fairly accurate even when new cases are imperfectly measured (11-13). If *R* is significantly larger than 1 for a sustained period, we identify that time period as an upward period, and inversely, If R is significantly smaller than 1 for a sustained period, we identify a downward period.

Based on the working definition and the proposed operationalization, we used public data on *R* to identify 179 countries that experienced some COVID-19 epidemic waves from 29^th^ March 2020 to 26^th^ Jan 2021. This working definition and operationalization, as we demonstrate, is straightforward yet can be customized or subject to robustness checks. We hope to offer a stepstone towards a common understanding of what constitutes an epidemic wave and enables us to be more effective in combating the continuing pandemic.

## Calibrating a working operationalization for COVID-19

We now describe the procedure to calibrate a working operationalization for COVID-19 (details in the *Supplement*). We illustrate one possible calibration of a working definition of epidemic waves of COVID-19 based on the COVID-19 contexts, especially considering the temporality of transmission and the cyclicity of infection case reporting. An individual can exhibit symptoms 11 days after the initial infection, with a confidence interval of 8 to 15 days (14). That is, the reporting of Covid-19 cases of people infected on day *t* will appear between day *t* + 8 to day *t* + 15. This variance in the temporal lag implies a potential covariance among R_*t*_ estimates within 7 days from each other. Therefore, it is a reasonable calibration to set *J*=7. Having *J* = 7 implies that at *n* must be at least 7. The choice of *n* should capture consecutive days to allow at least a two-chain transmission process to occur to model the sequential propagation of the epidemics in a region. The serial interval of one sequential propagation is estimated to be between 4 and 6 days (15), and we take 6 days, the upper limit, for prudence, and therefore *n* needs to be at least *7+6* = 13 days to capture a two-chain propagation in epidemic transmissions. Another critical consideration is the weekly cyclicity in COVID-19 infection cases seasonality (e.g., fewer cases on Sundays in many countries). To account for the weekly cyclicity, we try to set *n* = 14. An upward (downward) period refers to a period of at least n (=14) consecutive days when *R*_*t*_ is bigger (smaller) than 1.

The pandemic data used are the daily estimations of *R*_*t*_ that can be downloaded freely from their website (16) for each country when they have had at least 100 infections or more. The serial interval option used is the default option at 5 (the public R data contains several option intervals). We have incorporated results up to January 26th, 2020.

### Number and duration of upward and downward waves

The definition of a wave necessarily implies a period in which the number of cases goes up. After an upward, it is both possible for the cases to stabilize or go down. Table 1 reports the total amount of upward periods from March 29, 2020 to January 26, 2021 of 179 countries. There are 270 upward periods in total. Most countries (73.2%) have had at least one upward periods. Specifically, the number of countries with 1, 2, 3, 4, 5 upward periods are 45, 52, 20, 9 and 5 respectively. There are 48 countries without any upward period, and such countries are mostly small countries, island countries, or undeveloped countries that testing might be lacking. There are a total of 37 downward periods. Most countries (85.4%) had no downward period so far. Specifically, the number of countries with 1, 2, 3 downward periods are 19, 6, and 2, respectively. The number of downward periods does not match the number of upwards periods because many countries still had not had success in decreasing their cases for a sustained period of time to constitute a downward period by early 2021.

**Table 1.**
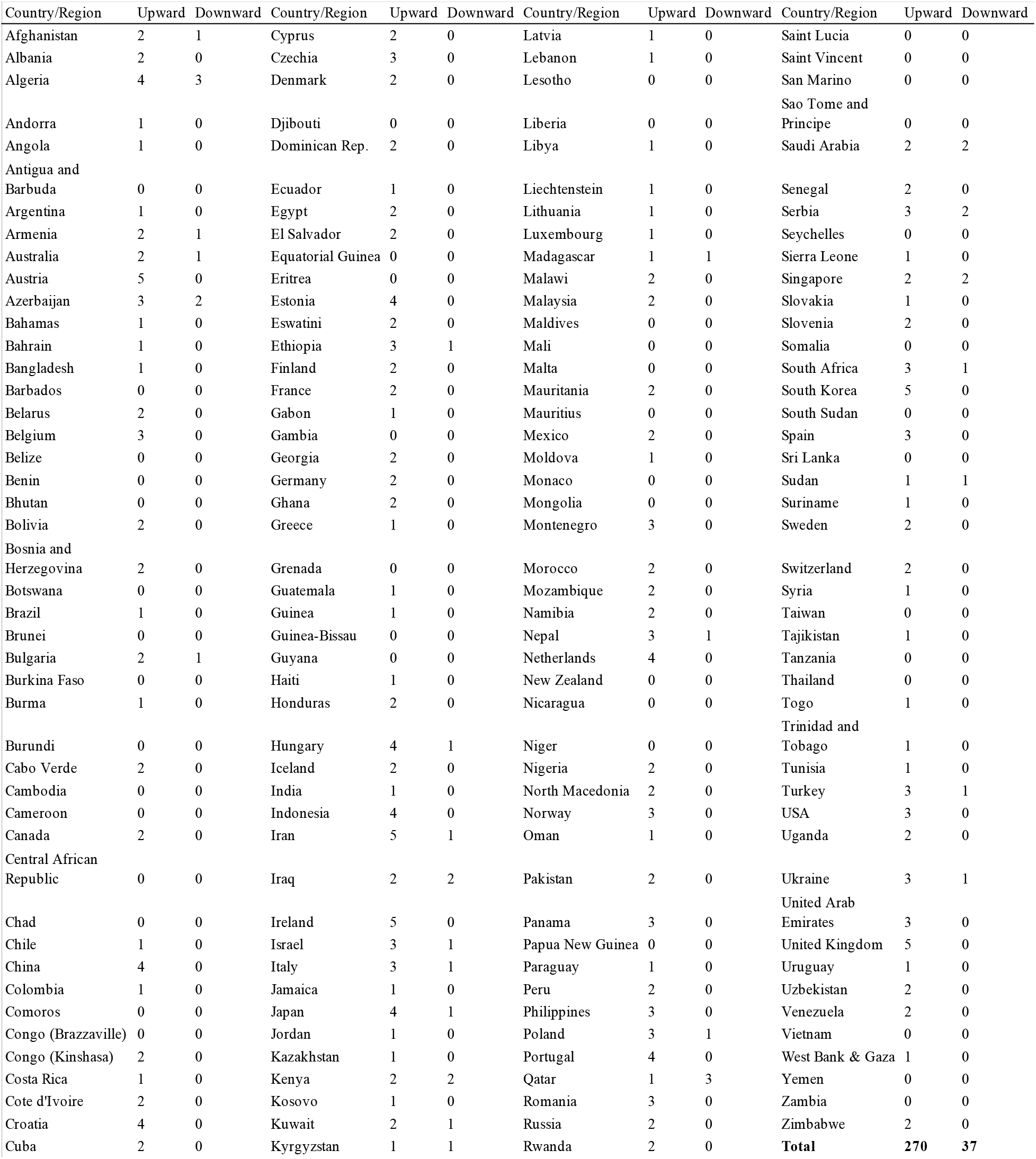
Number of upwards and downwards periods across countries

| Country/Region | Upward | Downward | Country/Region | Upward | Downward | Country/Region | Upward | Downward | Country/Region | Upward | Downward |
| --- | --- | --- | --- | --- | --- | --- | --- | --- | --- | --- | --- |
| Afghanistan | 2 | 1 | Cyprus | 2 | 0 | Latvia | 1 | 0 | Saint Lucia | 0 | 0 |
| Albania | 2 | 0 | Czechia | 3 | 0 | Lebanon | 1 | 0 | Saint Vincent | 0 | 0 |
| Algeria | 4 | 3 | Denmark | 2 | 0 | Lesotho | 0 | 0 | San Marino | 0 | 0 |
| Andorra | 1 | 0 | Djibouti | 0 | 0 | Liberia | 0 | 0 | Sao Tome and Principe | 0 | 0 |
| Angola | 1 | 0 | Dominican Rep. | 2 | 0 | Libya | 1 | 0 | Saudi Arabia | 2 | 2 |
| Antigua and Barbuda | 0 | 0 | Ecuador | 1 | 0 | Liechtenstein | 1 | 0 | Senegal | 2 | 0 |
| Argentina | 1 | 0 | Egypt | 2 | 0 | Lithuania | 1 | 0 | Serbia | 3 | 2 |
| Armenia | 2 | 1 | El Salvador | 2 | 0 | Luxembourg | 1 | 0 | Seychelles | 0 | 0 |
| Australia | 2 | 1 | Equatorial Guinea | 0 | 0 | Madagascar | 1 | 1 | Sierra Leone | 1 | 0 |
| Austria | 5 | 0 | Eritrea | 0 | 0 | Malawi | 2 | 0 | Singapore | 2 | 2 |
| Azerbaijan | 3 | 2 | Estonia | 4 | 0 | Malaysia | 2 | 0 | Slovakia | 1 | 0 |
| Bahamas | 1 | 0 | Eswatini | 2 | 0 | Maldives | 0 | 0 | Slovenia | 2 | 0 |
| Bahrain | 1 | 0 | Ethiopia | 3 | 1 | Mali | 0 | 0 | Somalia | 0 | 0 |
| Bangladesh | 1 | 0 | Finland | 2 | 0 | Malta | 0 | 0 | South Africa | 3 | 1 |
| Barbados | 0 | 0 | France | 2 | 0 | Mauritania | 2 | 0 | South Korea | 5 | 0 |
| Belarus | 2 | 0 | Gabon | 1 | 0 | Mauritius | 0 | 0 | South Sudan | 0 | 0 |
| Belgium | 3 | 0 | Gambia | 0 | 0 | Mexico | 2 | 0 | Spain | 3 | 0 |
| Belize | 0 | 0 | Georgia | 2 | 0 | Moldova | 1 | 0 | Sri Lanka | 0 | 0 |
| Benin | 0 | 0 | Germany | 2 | 0 | Monaco | 0 | 0 | Sudan | 1 | 1 |
| Bhutan | 0 | 0 | Ghana | 2 | 0 | Mongolia | 0 | 0 | Suriname | 1 | 0 |
| Bolivia | 2 | 0 | Greece | 1 | 0 | Montenegro | 3 | 0 | Sweden | 2 | 0 |
| Bosnia and Herzegovina | 2 | 0 | Grenada | 0 | 0 | Morocco | 2 | 0 | Switzerland | 2 | 0 |
| Botswana | 0 | 0 | Guatemala | 1 | 0 | Mozambique | 2 | 0 | Syria | 1 | 0 |
| Brazil | 1 | 0 | Guinea | 1 | 0 | Namibia | 2 | 0 | Taiwan | 0 | 0 |
| Brunei | 0 | 0 | Guinea-Bissau | 0 | 0 | Nepal | 3 | 1 | Tajikistan | 1 | 0 |
| Bulgaria | 2 | 1 | Guyana | 0 | 0 | Netherlands | 4 | 0 | Tanzania | 0 | 0 |
| Burkina Faso | 0 | 0 | Haiti | 1 | 0 | New Zealand | 0 | 0 | Thailand | 0 | 0 |
| Burma | 1 | 0 | Honduras | 2 | 0 | Nicaragua | 0 | 0 | Togo | 1 | 0 |
| Burundi | 0 | 0 | Hungary | 4 | 1 | Niger | 0 | 0 | Trinidad and Tobago | 1 | 0 |
| Cabo Verde | 2 | 0 | Iceland | 2 | 0 | Nigeria | 2 | 0 | Tunisia | 1 | 0 |
| Cambodia | 0 | 0 | India | 1 | 0 | North Macedonia | 2 | 0 | Turkey | 3 | 1 |
| Cameroon | 0 | 0 | Indonesia | 4 | 0 | Norway | 3 | 0 | USA | 3 | 0 |
| Canada | 2 | 0 | Iran | 5 | 1 | Oman | 1 | 0 | Uganda | 2 | 0 |
| Central African Republic | 0 | 0 | Iraq | 2 | 2 | Pakistan | 2 | 0 | Ukraine | 3 | 1 |
| Chad | 0 | 0 | Ireland | 5 | 0 | Panama | 3 | 0 | United Arab Emirates | 3 | 0 |
| Chile | 1 | 0 | Israel | 3 | 1 | Papua New Guinea | 0 | 0 | United Kingdom | 5 | 0 |
| China | 4 | 0 | Italy | 3 | 1 | Paraguay | 1 | 0 | Uruguay | 1 | 0 |
| Colombia | 1 | 0 | Jamaica | 1 | 0 | Peru | 2 | 0 | Uzbekistan | 2 | 0 |
| Comoros | 0 | 0 | Japan | 4 | 1 | Philippines | 3 | 0 | Venezuela | 2 | 0 |
| Congo (Brazzaville) | 0 | 0 | Jordan | 1 | 0 | Poland | 3 | 1 | Vietnam | 0 | 0 |
| Congo (Kinshasa) | 2 | 0 | Kazakhstan | 1 | 0 | Portugal | 4 | 0 | West Bank & Gaza | 1 | 0 |
| Costa Rica | 1 | 0 | Kenya | 2 | 2 | Qatar | 1 | 3 | Yemen | 0 | 0 |
| Cote d'Ivoire | 2 | 0 | Kosovo | 1 | 0 | Romania | 3 | 0 | Zambia | 0 | 0 |
| Croatia | 4 | 0 | Kuwait | 2 | 1 | Russia | 2 | 0 | Zimbabwe | 2 | 0 |
| Cuba | 2 | 0 | Kyrgyzstan | 1 | 1 | Rwanda | 2 | 0 | **Total** | **270** | **37** |

The duration of upward periods varied too. The average upward period’s length is 61.7 days (s.d.= 47.0), and three quarters of them lasted less than 31 days. The minimum and maximum upward were 15 and 302 days, respectively. The distribution is skewed positively with a value of 2.4, as illustrated in Figure 1a, which shows the distribution of the duration of the 270 upward periods. The duration of downward periods averaged 33.6 days (s.d.= 17.0). The distribution is also skewed positively, with a value of 3.1, as illustrated in Figure 1b.

**Figure 1a.**
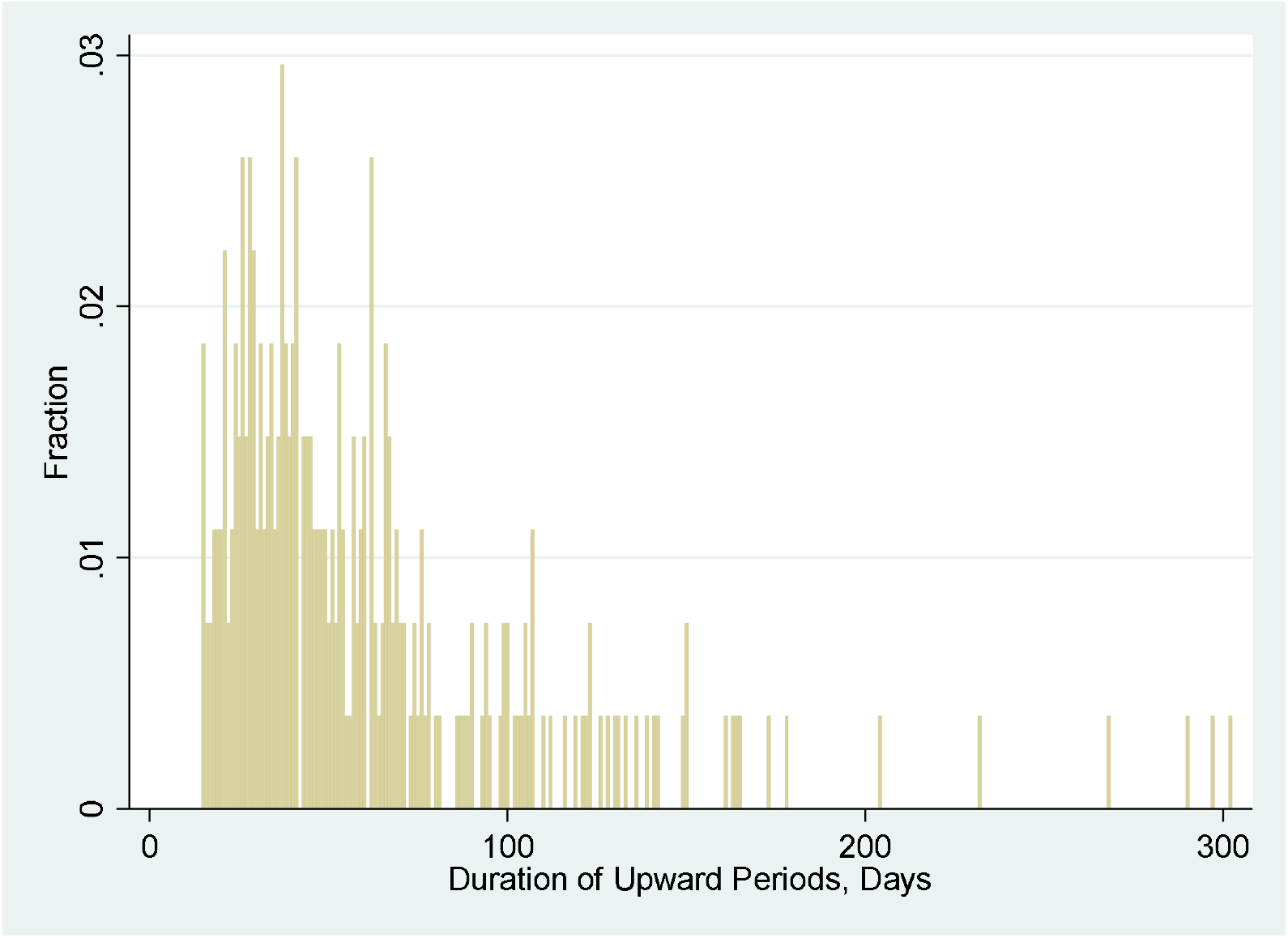
The distributions of upward period duration

**Figure 1b.**
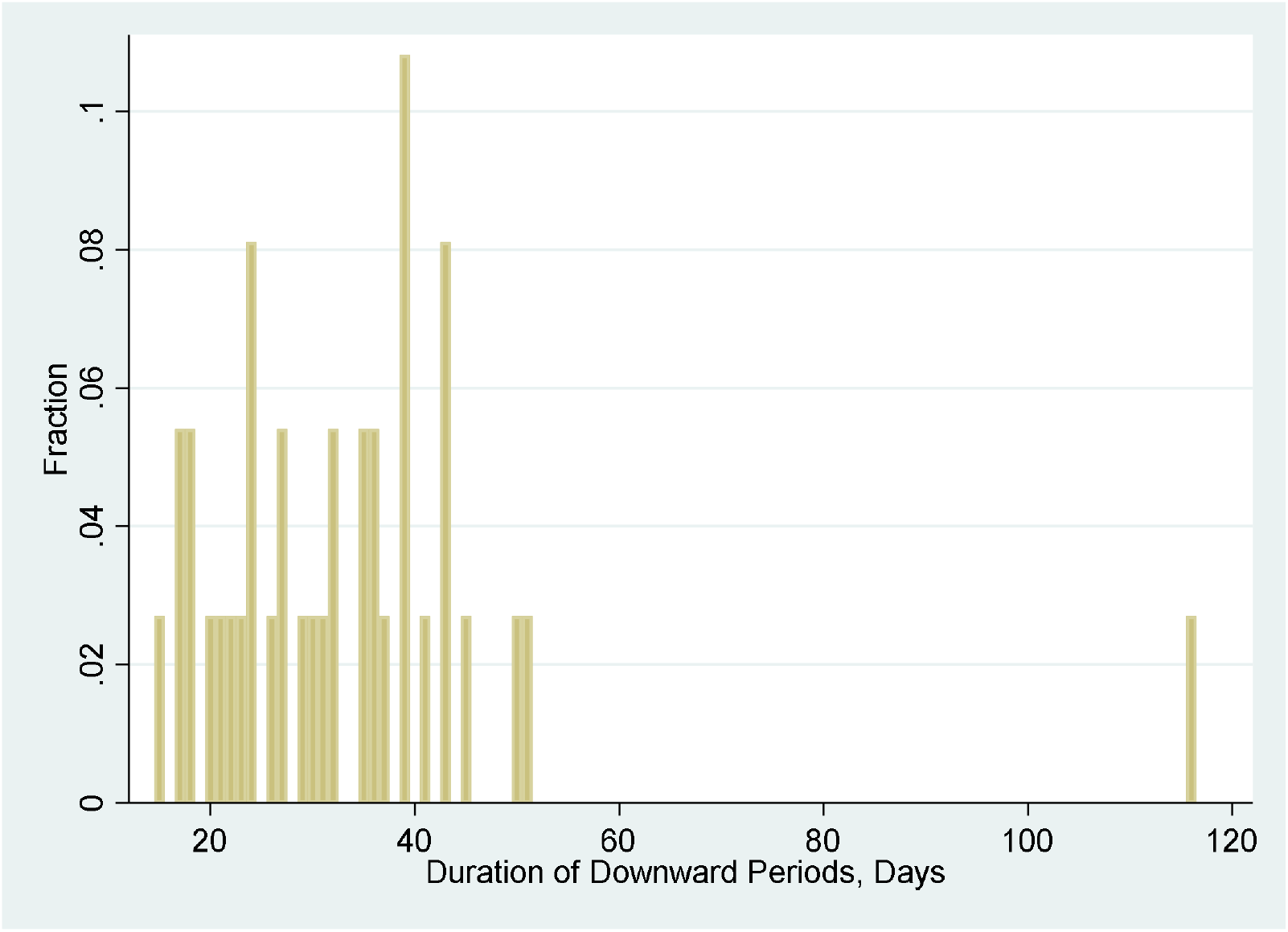
The distributions of downward period duration

## DISCUSSION

This paper offers a working definition of epidemic waves that can characterize the ongoing COVID pandemic in rea-time. The data used to operationalize the epidemic wave are publicly available (e.g., the website of John Hopkins COVID-19 statistics) not only at the country-level but also at the more granular regional levels (e.g., the USA states) to characterize epidemic waves at different levels. It is also possible to include the upward and downward periods of waves in such websites to enable a common ground for public and the policymakers to understand the ongoing epidemics.

First, it is essential to characterize waves, and our working definition and an operationalization provides a common ground for health communication, as to date such a common ground is lacking among different world leaders and healthcare experts. Our characterization approach consists of a statistical criterion that can be applied in real-time, so that we are not subject to various *post hoc* interpretations of waves, as it happened in the Spanish flu pandemic. The characterization of waves can help policymakers to make more informed decisions on resource mobilization and distribution over time as the COVID-19 epidemic evolves. Reliable descriptions are essential for estimating the true impact of COVID-19 and the control of COVID-19 across countries over time. “All of this requires collection of clear, uniform and comparable” characterization of the epidemic in real-time, and we hope our working definition of an epidemic wave offers a springboard in that direction (17).

Second, our findings that most countries (e.g., USA and UK) have had more upward periods than downward periods in the year 2020 suggest that these countries had not decreased the infection for a sustained period of time. The containment strategies in many countries (especially developed countries) have broken the upward periods (i.e., upward, stagnant, and then upward) but did not result in a sustained period of time and hence may effectively just delayed the epidemic and could have wasted the window of opportunities. Indeed, many countries and regions declared success in their containment strategies in mid-2020 while in fact they did not decrease the infection for a sustained period of time. Our characterization of epidemic waves can help to examine the effectiveness of interventions from the identification of downward periods as well as the triggers of upward periods across regions to prevent the occurrence of subsequent waves.

While it is hard to compare the daily or total COVID-19 deaths and cases between countries due to their different sizes, waves capture the changes (upward or downward) within a country and hence present an alternative approach to assess the fluctuation of epidemics in a country or a region. The working definition of epidemic waves can be universally applied across countries/regions, hence offering an alternative way that is more comparable, given that “epidemiology is built on the idea of studying differences between populations. Much of what we have learnt about the causes of disease has had its origins in comparisons of countries” (18). As countries adopt different approaches over time to deal with the COVID-19, it offers some possibilities to learn what works best for controlling COVID-19, given the lack of real randomized control experiments in epidemiology. Epidemiologists call for “more thoughtful and thorough analyses of country differences, … remain probably the most important and most valid evidence for informing COVID-19 policy in real-time” (18). Specifically, the identification of the upward and downward periods (counts and durations) in various countries may be matched with the containment strategies in those countries to help identify effective strategies to curtail the wave.

### Limitation and future research

Our approach is limited by the following points. First, COVID-19 daily cases may be misreported, and reporting delays and inconsistent practices for defining and testing COVID-19 cases may also influence the accuracy of daily cases, which are the input to calculate the reproduction rate and hence the characterization of waves. To reduce the noise and errors in daily reporting, our definition picks the average of the past 14 days. However, the average over 14-day samples does come at a trade-off that a sharp to exceptionally low values within a couple of days may not be statistically significant. Our choice of the parameter offers one possible illustration of the calibration of a working definition of epidemic waves, and other calibrations are possible. Our working definition characterizes waves by upward periods and downward periods, the fundamental elements of waves. We notice many broken waves (i.e., an upward period, a break, an upward period), so epidemic waves may not follow a symmetrical pattern of an upward period followed by a downward period in some waves. Moreover, as the COVID-19 crisis still continues and worsens in many parts of the world, our findings remain incomplete and need to be updated as new data arrive.

## Conclusion

A wave of COVID-19 cannot be declared arbitrarily by any individuals without a common definition. The first questions we need to understand are: ‘what do we mean by epidemic waves, then where do waves occur and not occur, and what can we learn from this?’ We hope that our working definition and operationalization offers a stepping stone to better understand the epidemics to help with containing the continuing COVID-19 crisis.

## Data Availability

it used public data

## Acknowledgments

Nil.

## Author contributions

- Conceptualization: SXZ.
- Methodology: FAM.
- Investigation: SXZ, FAM, FG
- Visualization: FAM, FG
- Funding acquisition: nil
- Project administration: SXZ
- Supervision: SXZ
- Writing – original draft: SXZ, FAM, FG
- Writing – review & editing: SXZ, FG. FAM

## Data and materials availability

All data, code, and materials used in the analysis are available online publicly and can be requested from the corresponding author.

## SUPPLEMENT

To characterize an epidemic wave of having at least an upward and/or downward periods for a sustained period of time in a country, we employ R_*t*_, a time series estimates of the reproduction number (11). If the average reproduction rate in the last *n* days is above 1, there is evidence of a sustained upward period. Similarly, if the average reproduction rate in the last *n* days is below 1, there is evidence of a sustained downward period. Formally, we test the null hypothesis 1 and alternative hypothesis 2, where 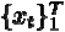 denotes the daily estimates of R_*t*_ at any moment *T* in the last *n* number of days, i.e. *t* = 1,…,*n*. Equation 3 refers to the sample average of R_*t*_ estimates during the past n days.

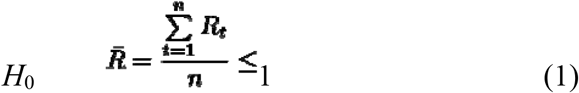

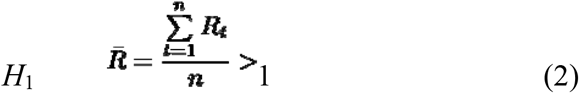

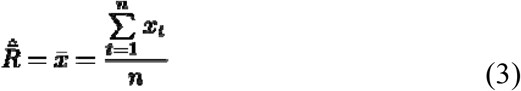

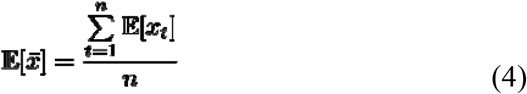

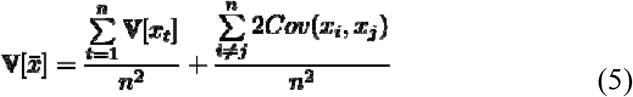

Equations 4 and 5 show the formula for the expectations and variance for any simple average estimators, in this case, the simple average of R_*t*_ estimates of the past *n* days. The expectation of average *R*^-^_*t*_ is calculated by taking the average of the estimate of the mean or median of the posterior distribution of each daily estimation. Because the estimates of R_*t*_ use a Kalman filter estimation (11), where both the transition and state equations have normally distributed error terms, the posterior distribution of each of the estimates will rapidly converge to a normal distribution, and the weighted sum of normally distributed variables, ^-^*x*, is also normally distributed. The variance, however, depends on whether the estimates were completely independent between themselves. If so, the second term in equation 5 would be zero and calculation of V[^-^*x*] would be the sum of the variance of each estimate in the sample, divided by *n*^*2*^. However, the covariance among some estimates may not be zero, as it is likely to have positive covariance in adjacent dates in a time series. Since it is impossible to know the value of these covariance terms in real-time in reality, we use an upper bound for the covariance of adjacent variables in the second term based on the statistical property that the covariance between two variables can never be bigger than the biggest variance of both variables as the upper bound for the variance, described in equation 5.

### The number and duration of upward and downward waves by regions and country groups

We break down the count and duration of upward and downward first by developed and developing (including undeveloped) economies. Developed countries have experienced a similar number of upward periods to developing countries (133 times vs. 137 times) but shorter upward periods (52.9 days vs. 70.2 days; d=-17.3 days) than developing countries. Moreover, developed countries have experienced a similar number of downward periods to developing countries (19 times vs. 18 times) but shorter downward periods (30.5 days vs. 36.9 days; d=-6.4 days) than developing countries. Most of the developed economies have gone through more than one upward periods, though not always followed by a downward one (65.1% that have at least one upward period have no downwards). For example, the US has suffered three upward periods, but not any downward period; Italy has had three upward periods but only one downward period. Many developed economies have had upward periods, followed by some effort to contain the upward trend, yet the effort has mostly tempered the reproduction to stabilize it rather than being able to lead to a sustained downward period. Such a characterization can help policymakers and healthcare organizations to better understand the typical patterns by not only economic development but also by other attributes (e.g., cultural, institutional, populational factors), for better planning, resource allocation and policy decisions in advance. For instance, health authorities can better plan the temporal demand for medical resources, whereas economic policymakers can be more informed in drafting policies.

### Correlation between upward/downward counts/duration and other country-level statistics

We examine how the duration and count of upward and downward periods correlated with each other and other key country-level statistics (Table 2). The duration of upward periods and the duration of downward periods are negatively correlated (r=-0.27, p=.00), indicating countries with longer upward periods tend to have shorter downward periods. The number of upward and the duration of downward period in a country are also positively correlated (r=0.27, p=.00), indicating countries with more upward periods tend to have more downward periods.

**Table 2.** Correlations matrix: the number/duration of upward/downward periods and key country-level indicators (number of countries: *n* = 177)

|  | 1 | 2 | 3 | 4 | 5 | 6 | 7 | 8 | 9 | 10 | 11 | 12 | 13 | 14 |
| --- | --- | --- | --- | --- | --- | --- | --- | --- | --- | --- | --- | --- | --- | --- |
| **1** Upward count | 1 |  |  |  |  |  |  |  |  |  |  |  |  |  |
| **2** Downward count | 0.268<br>*0.000* | 1 |  |  |  |  |  |  |  |  |  |  |  |  |
| **3** Upward duration | -0.524<br>*0.000* | -0.217<br>*0.000* | 1 |  |  |  |  |  |  |  |  |  |  |  |
| **4** Downward duration | -0.172<br>*0.000* | -0.067<br>*0.000* | -0.236<br>*0.000* | 1 |  |  |  |  |  |  |  |  |  |  |
| **5** Stringency index | 0.264<br>*0.000* | -0.052<br>*0.000* | -0.022<br>*0.000* | -0.323<br>*0.000* | 1 |  |  |  |  |  |  |  |  |  |
| **6** GDP per capita | 0.276<br>*0.000* | 0.231<br>*0.000* | -0.090<br>*0.000* | -0.143<br>*0.000* | 0.206<br>*0.000* | 1 |  |  |  |  |  |  |  |  |
| **7** Total cases | 0.030<br>*0.000* | 0.001<br>*0.885* | 0.038<br>*0.000* | -0.208<br>*0.000* | 0.235<br>*0.000* | 0.045<br>*0.000* | 1 |  |  |  |  |  |  |  |
| **8** Total cases<br>per million | 0.276<br>*0.000* | 0.027<br>*0.000* | 0.028<br>*0.000* | -0.346<br>*0.000* | 0.346<br>*0.000* | 0.486<br>*0.000* | 0.165<br>*0.000* | 1 |  |  |  |  |  |  |
| **9** Total death | 0.054<br>*0.000* | -0.007<br>*0.145* | -0.001<br>*0.882* | -0.208<br>*0.000* | 0.257<br>*0.000* | 0.040<br>*0.000* | 0.905<br>*0.000* | 0.162<br>*0.000* | 1 |  |  |  |  |  |
| **10** Total death<br>per million | 0.267<br>*0.000* | -0.043<br>*0.000* | -0.010<br>*0.049* | -0.370<br>*0.000* | 0.369<br>*0.000* | 0.280<br>*0.000* | 0.268<br>*0.000* | 0.821<br>*0.000* | 0.371<br>*0.000* | 1 |  |  |  |  |
| **11** Human development<br>Index (HDI) | 0.396<br>*0.000* | 0.107<br>*0.000* | -0.135<br>*0.000* | -0.240<br>*0.000* | 0.429<br>*0.000* | 0.751<br>*0.000* | 0.162<br>*0.000* | 0.597<br>*0.000* | 0.182<br>*0.000* | 0.528<br>*0.000* | 1 |  |  |  |
| **12** Population density | -0.052<br>*0.000* | 0.073<br>*0.000* | -0.041<br>*0.000* | 0.032<br>*0.003* | -0.037<br>*0.000* | 0.293<br>*0.000* | -0.031<br>*0.000* | 0.035<br>*0.000* | -0.043<br>*0.000* | -0.058<br>*0.000* | 0.141<br>*0.000* | 1 |  |  |
| **13** Positive COVID-19 rate | 0.095<br>*0.000* | -0.132<br>*0.000* | 0.076<br>*0.000* | 0.036<br>*0.005* | 0.006<br>*0.381* | -0.357<br>*0.000* | -0.001<br>*0.880* | 0.105<br>*0.000* | 0.214<br>*0.000* | 0.295<br>*0.000* | -0.264<br>*0.000* | -0.186<br>*0.000* | 1 |  |
| **14** Total tests | -0.054<br>*0.000* | -0.032<br>*0.000* | 0.069<br>*0.000* | -0.141<br>*0.000* | 0.108<br>*0.000* | -0.013<br>*0.049* | 0.949<br>*0.000* | 0.005<br>*0.436* | 0.713<br>*0.000* | 0.039<br>*0.000* | 0.034<br>*0.000* | 0.061<br>*0.000* | -0.168<br>*0.000* | 1 |
| **15** Total tests<br>per thousand | 0.058<br>*0.000* | -0.073<br>*0.000* | 0.120<br>*0.000* | -0.233<br>*0.000* | 0.098<br>*0.000* | 0.624<br>*0.000* | -0.031<br>*0.000* | 0.544<br>*0.000* | -0.071<br>*0.000* | 0.157<br>*0.000* | 0.503<br>*0.000* | 0.186<br>*0.000* | -0.332<br>*0.000* | 0.081<br>*0.000* |

Countries with higher GDP per capita tend to have shorter duration of upward periods (r= - 0.09, p=.00) and downward periods (r= -.14, p=.00) yet more number of upward periods (r= .28, p=.00) and downward periods (r= .32, p=.00). Similarly, countries with higher Human Development Index (HDI) tend to have shorter duration of upward periods (r= -.14, p=.00) and downward periods (r= -.24, p=.00) yet more number of upward periods (r= .40, p=.00) and downward periods (r= .11, p=.00). Lastly we examine the correlation with the stringency index, which reveals that countries with more stringent measures on COVID-19 have shorter duration of upward periods (r= -.02, p=.00) and downward periods (r= -.32, p=.00) and more upward periods (r= .26, p=.00) and fewer downward periods (r=-.05, p=.00).

Such patterns can inform policymakers and healthcare organizations. For instance, knowing the pattern that longer upward periods are usually followed by longer downward periods has direct implications for planning and resource allocation decisions on medical supplies, personnel, infrastructure, etc. Such information is crucial not only for healthcare organizations and policies but also for social and economic policies, as the COVID-19 situation changers the social-economic policies that aim to cope with the epidemic situations.

